# Factors influencing COVID-19 vaccination uptake in an elderly sample in Poland

**DOI:** 10.1101/2021.03.21.21254047

**Authors:** Marta Malesza, Magdalena Bozym

## Abstract

**Backgrounds:** This research represents an investigation into potential predictors for the uptake of the COVID-19 vaccination in Poland, following the instigation of policies to encourage the over-seventies to be vaccinated.

**Methods:** Individuals participated in cross-sectional structured interviews. 1427 respondents were questioned for determining vaccination uptake, revealing attitudes regarding vaccination, where information was sourced from, health status and behavior, demographics and socio-economic profiles.

**Results:** Selected predictors for acceptance of the vaccination were: being talked through the importance of the vaccination and potential side-effects by a medical professional; sharing living space with others; having a high ranking occupation; suffering from chronic illnesses; being able to access medical services by driving or walking rather than using public transport or relying on others. Those who opted not to be vaccinated most frequently justify their decision by saying that they were concerned about the efficacy of the vaccine or that they were worried about side-effects.

**Conclusions:** It appears that the current nationwide campaign has successfully raised awareness regarding the vaccine, but this research indicates that a more information-based campaign, focusing on evidence of the vaccine’s efficacy and the non-serious nature of all side-effects, could lead to improved uptake of the COVID-19 vaccine.

## Introduction

In late 2019 a novel coronavirus variant (COVID-19) appeared in Wuhan province (China); this virus swiftly spread globally. The World Health Organization (WHO) classified the outbreak as a Public Health Emergency of International Concern on January 30, 2020; a pandemic was declared on March 11, 2020 [1]. By January 31, 2021, 2,216,363 residents of Poland had contracted coronavirus, 56,945 of whom had died [2]. In accordance with WHO recommendations, Poland initiated pandemic preparedness planning, the central strand of which was a vaccination program [3, 4]. A mass vaccination program began in Poland in the last weeks of 2020.

There is a significant lack of research regarding the elements that influence the uptake of COVID-19 vaccines now that they are publicly available [5,6]. The majority of past research regarding vaccine uptake was undertaken prior to the pandemic when vaccines against COVID-19 did not exist; other research was limited in its research cohort, e.g. focusing only on healthcare professionals [7, 8, 9, 10]. This means that the extant literature may not represent an accurate indicator of the probability of vaccine acceptance due to the fact that the ways in which the general public perceives the pandemic and potential vaccines will change with changes in circumstances. In addition, the attitudes of those working in healthcare and the general public are very different: healthcare workers have much higher risks of contracting the virus due to their employment; they may also be more informed about the virus and vaccines because of their work. Research has demonstrated that healthcare workers in the emergency sector and those who have assumed additional duties as a result of the pandemic have a greater likelihood of accepting a vaccination against illness, as do those who have demonstrated a positive attitude regarding prophylactic measures against illness [10, 11].

COVID-19 is a particular hazard for elderly individuals [1]. Because of this, the Polish Minister of Health issued a recommendation that every individual aged 70+ should be encouraged to receive the COVID-19 vaccination [3]. In accordance with the Ministry’s recommendation, vaccinations were offered free of charge and a national campaign was undertaken, encouraging every citizen to accept a vaccination. It is essential that a full understanding is developed of the elements that influence vaccination uptake so that future advertising initiatives and government interventions can be better targeted. Vaccination programs are reliant for efficacy on widespread acceptance; this is the case even with extremely effective vaccines. This makes it paramount that we should have an understanding of the elements influencing an individual’s acceptance or otherwise of a vaccine, so that public health strategy for pandemics can be effectively developed and implemented [3, 4].

Past research has demonstrated that when elderly people who have not been given a flu vaccine it is generally because they have refused the vaccine, not because it has not been offered, although no assessment was made of why they refused [12]. For patients at high risk, the most frequent reasons for not being vaccinated were that vaccines had not been widely enough publicized or offered [13] and that potential patients held misconceptions about the vaccine’s efficacy and/or potential side-effects [14]. Some research [15] has indicated that amongst those elderly people still living in their community, high-risk individuals refused vaccination generally because they did not understand the level of risk involved and were not sufficiently advised by qualified healthcare personnel. Other research amongst the same type of cohort found that vaccine uptake predictors were belief in the vaccine’s effectiveness, experience of being vaccinated previously, and a lack of concern regarding side-effects [16]. In other research, it was shown that the primary predictors for refusing vaccines were patients who did not have a high risk classification, who believed they enjoyed robust health, who were not advised by healthcare professionals, and who viewed vaccines negatively in terms of safety and efficiency [17].

Qualitative research undertaken with elderly participants has indicated that being skeptical about the worth of a vaccine and concerns about potential side-effects had a greater influence than not being provided with sufficient advice from medical experts was the primary reason for non-vaccination [13, 15, 18]. It has been suggested that because the COVID-19 pandemic is so serious, patient character could be more influential regarding uptake than the level of medical advice received, because during the pandemic any eligible patients will have been advised by their general practitioner (GP) to have the vaccination. Therefore this research has examined COVID-19 vaccination uptake amongst Poland’s elderly population in order to achieve a better understanding of what influences uptake of the COVID-19 vaccine.

### Methodology

#### Research cohort

Poland’s six principal geographic regions encompass 16 administrative units (*voivodeships*), divided along on historic, cultural, economic, and geographic lines, each of which is subdivided into counties. A stratified sampling method was employed, and recruiters responsible for data collection were sent to randomly-chosen cities. Sampling of counties within voivodeships, whereby two counties from each voivodeships (total = 32 counties) were chosen. 3200 adults were spoken to in a variety of locations, e.g. around churches and in shopping centers, in 32 Polish cities that were selected, in January/ February 2021 and given an invitation to engage in a structured interview. From 3200 individuals, 2505 agreed to this orally once the research had been explained (78.3% response rate), with 1427 meeting the inclusion criteria (being aged 70+ and living amongst the general public, i.e. not in a care/nursing home; being vaccinated or denying vaccination against COVID-19). 708 participants were female (49.6%) and 719 were male (50.4%). Oral informed consent to participate in this study was given by all respondents. This research was granted approval by the appropriate ethics committee.

### Interview

Structured interviews were created for determining vaccination uptake, revealing attitudes regarding vaccination, where information was sourced from, health status and behavior, demographics and socio-economic profiles.

### Vaccination uptake

Respondents were questioned as to whether they had already been vaccinated against COVID-19, and if so when; those who had not been vaccinated were asked if they intended to accept a vaccine once it was offered (i.e., had they registered to be vaccinated)^1^. This form of self-reporting has been demonstrated to be extremely accurate in recording vaccination behaviors for the elderly [19].

### Demographics, socio-economic status, and accessibility

Respondents were asked their age group (70-79 or 80+) and if they lived by themselves or with other people. The gender of the respondent was noted. Respondents were questioned as to their current or previous occupations; if the respondent was a housewife, spousal occupation was noted. Occupation (self-reported) was classified in accordance with the National Statistics Social Economic Classification system (I = professional, II = managerial/technical, III = skilled, IV = partially skilled, V= unskilled); if the respondent had a partner, the higher status occupation of the two was used to identify the socio-economic status of their household. The rankings were used to create two categories of professional (Groups I and II) and nonprofessional (Groups III-V). Respondents were also questioned regarding their normal means of transport to their GP (walking, bus, personal automobile, automobile of friend or family, taxi, train, or GP visit at home) to show the accessibility of healthcare. Patients were placed in categories of either independent (walking or own automobile) or dependent (public transport, taxi, or lifts from friends or relatives).

### Health status/behavior

Respondents were questioned regarding smoking behaviors (always non-smoker, currently smoking, previously smoked) and alcohol intake (never, only on special occasions, one or two times a week, virtually every day). Respondents were placed in categories of smoker or non-smoker and occasional drinker or regular drinker. Respondents were also questioned as to whether they had any medical conditions that increased their risk around COVID-19, including kidney disease, heart disease, lung disease, liver disease, diabetes, and asthma (yes/no responses).

### Information sources

Respondents were questioned as to whether they had had an explanation as to why they should receive the vaccine from a medical professional and an explanation of the potential side-effects (both yes/no). They were also questioned as to whether they remembered seeing any advertising about COVID-19 vaccinations (yes/no) and, if they did, where (TV, radio, newspapers, leafleting, doctor’s surgery, other).

### Attitude towards COVID-19 vaccines

Respondents who indicated that they had taken the vaccine were questioned as to what influenced their decision (e.g., will to avoid sickness, need for being protected, advice from friends, advice from GP, reminder from GP, personal initiative). If they had not been given the vaccine and had no plans to accept it, they were questioned as to their reasons (family or friends having bad experiences of vaccines, personal bad experiences of vaccines, lack of awareness of vaccine, belief the vaccine was not effective, time considerations, being afraid of needles, concern regarding side-effects, medical reasons). Patients who had not been vaccinated were also questioned as to whether anything might change their mind about the vaccine (see Table 2).

### Data analysis

Logistic regression models underwent testing for determination of the correlation between vaccination uptake and attitudes, level of information, socio-economic status, demographics, and health status. Univariate predictors were then added to a multiple logistical regression for determining possible independent predictors of vaccine uptake.

## Results

### Attitudes towards COVID-19 vaccination

62.7% (895/1427) of respondents stated that they had been given the COVID-19 vaccination against COVID-19 while 37.3% (532/1427) of participants said they were unwilling to get vaccinated against COVID-19 (Table 1). The participants’ explanations for wanting or not to be vaccinated are presented in Table 2. The main reason to accept vaccination was ‘‘self-protection’’ (90.6%). Over 69% of respondents wanted to protect their close relatives by getting vaccinated. Almost half of the study participants (44.2%) believed that the vaccine could stop the coronavirus outbreak. 36.0% of respondents said that vaccines are safe and 33.2% of individuals stated that vaccines have no side effects. Additional important influences on accepting a vaccination were receiving a reminder from a doctor (67.7%), receiving advice from a medical professional (65.0%) or from a friend (49.9%). Only 17.8% invoked that getting vaccinated was ‘‘a civic duty’’ (Table 2).

**Table 1.**
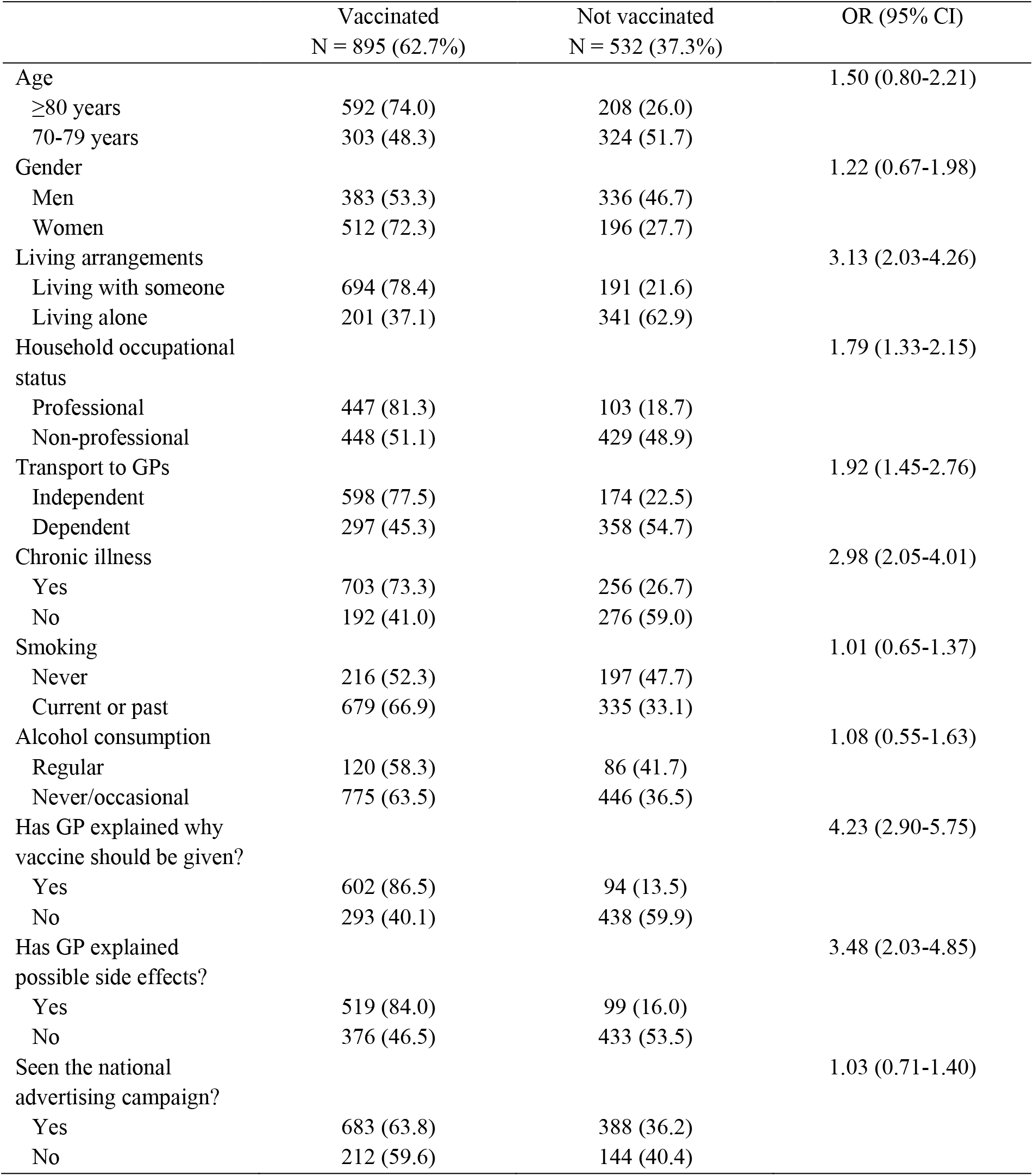
Logistic regression models of predictors of reported influenza vaccination uptake

**Table 2.** Reasons for acceptability or not of COVID-19 pandemic vaccination in the Polish elderly population.

| **Main reason(s) for acceptability of pandemic vaccination (N = 895)** | **% (N <sup>1</sup>)** |
| --- | --- |
| Protecting myself to avoid sickness | 90.6% (811) |
| Protecting my close relatives | 69.3% (620) |
| Received a reminder from a doctor | 67.7% (606) |
| Vaccination is recommended by health officials (GP) | 65.0% (582) |
| Vaccination was recommended by friends | 49.9% (447) |
| The vaccine will stop the outbreak | 44.2% (396) |
| Vaccines are safe | 36.0% (322) |
| Vaccines have no side effects | 33.2% (297) |
| Getting vaccinated is convenient and quick | 22.6% (202) |
| Getting vaccinated is a civic duty | 17.8% (159) |
| **Main reason(s) for non-acceptability of pandemic vaccination (N = 532)** | **% (N <sup>1</sup>)** |
| Vaccines are not safe enough | 91.4% (486) |
| Vaccines have side effects | 89.7% (477) |
| COVID-19 is not a severe disease | 75.4% (401) |
| I don't want to be a guinea pig | 69.5% (370) |
| Vaccines are ineffective | 66.7% (355) |
| Family or friends had bad experiences of vaccines | 57.1% (304) |
| I have medical reasons to avoid vaccines | 53.0% (282) |
| I dislike the shots | 35.3% (188) |
| Getting vaccinated is inconvenient and too long | 30.8% (164) |
| Personal bad experiences of vaccines | 19.2% (102) |
| I don't know | 9.4% (50) |

Note: Any items could be selected and thus proportions do not add to 100%. Items were presented in a random order.
1 = where the answers were grouped as „definitely yes” and „probably yes”.

Among respondents who did not get vaccinated, the main reasons were concerns about vaccine safety and fear of vaccine side effects (answered by 91.4% and 89.7% respondents, respectively). Three-fourth of respondents (75.4%) stated that COVID-19 is not a severe disease. Also 69.5% of individuals said they do not want to be used as the subject of an experiment. Almost the same percent of people (66.7%) stated that vaccines are ineffective. Another important influences on not being vaccinated were medical conditions that contraindicated the vaccine (53.0%), or family/friends having had a bad experience of the vaccine (57.1%). 30.8% thought that getting vaccinated is inconvenient. 9.4% were not able to give any reason for not (Table 2).

Respondent percentages regarding vaccination by predictor variable is shown in Table 1 along with the odds ratio.

### Demographics, socio-economic status, accessibility

Respondents who lived alone had a lower likelihood of having received the vaccine than those who lived with others. Neither age nor gender was a significant predictor of vaccination. Respondents capable of independent private travel to their GP, those suffering from chronic illnesses, and those in the higher socio-economic groups, had a greater likelihood of being vaccinated.

### Information sources

Those who had been given an exclamation by medical professional of why they should be vaccinated and the potential side-effects were more likely to have accepted a vaccine. 75.1% of respondents stated that they had seen the nationwide campaign regarding COVID-19 vaccinations, but this was not a significant predictor of vaccine acceptance.

### Multiple logistic regression model for prediction of COVID-19 vaccination uptake

For a multiple logistic regression model with every significant univariate predictor entered, the most significant independent predictors of vaccine acceptance were being given an explanation by a medical professional as to why they should be vaccinated and sharing their habitation with others. Being able to travel independently to their GP, having chronic illnesses, and respondents’ socio-economic status were also predictors of vaccine acceptance. In this model, there was also a significant correlation between being given an explanation by a medical professional of potential side-effects and vaccine acceptance.

## Discussion

This research revealed that, for this elderly cohort, the COVID-19 vaccination program has been quite successful, with over a half of participants stating that they had been given the current COVID-19 vaccination. The main predictors for accepting the COVID-19 vaccine were sharing habitation with others, being able to get to the GP surgery independently, suffering from chronic illnesses, and being informed of the importance and side-effects of the vaccination by a medical professional. This implies that being given medical advice and individual patient characteristics are both important contributions to acceptance of COVID-19 vaccinations for elderly individuals. This went against the initial hypothesis that, since policy changed to encompass targeting of all 70+ citizens, being given medical advice would be less important as a vaccine acceptance predictor than individual patient characteristics. In the event, self-reported justifications for accepting the vaccine were primarily to do with medical encouragement. Most of those who had accepted a vaccination did so because they had received advice or a reminder from healthcare professionals.

Perceptions of having no impact on outcomes or, in other words, feelings of powerlessness [22] are often associated with beliefs in conspiracy theories [23]. Thus, it is possible that a pre-existing feeling of inability to take charge of one’s own wellbeing render individuals more susceptible to agreement with anti-vaccine theories, thus reducing willingness to be vaccinated. This would explain why we found that the most frequently cited driver for refusal to be vaccinated against COVID-19 was the concern it might not be safe. Research has shown that pandemics provoke inquiry into vaccine efficacy and safety on a global scale and that if scientific evidence is not put in the public domain to prove these matters, vaccination take-up rates will remain low in the general population [25].

The apparent success of national advertising campaigns and reminders from GPs is demonstrated by the fact that just a small percent of respondents had no awareness of the COVID-19 vaccination program. Nevertheless, those who did not accept the vaccine refused it principally due to misinformation regarding the effectiveness or safety of the vaccine and potential side-effects. This indicates that uptake could be further improved by providing more accurate information about the vaccine and ensuring all patients are invited to receive it. Subsequent public information campaigns should be focused on demonstrating that side-effects are not serious, that the vaccine works, and that the vaccine will not make people ill [7-10, 25-28].

Although the levels of respondents saying they had not been given the vaccine due to access problems were relatively low, this research is the first that indicates that individuals who live alone or who cannot travel independently to their GP had a lower likelihood of having had the vaccine. This indicates that individuals in such a position should be offered additional support and encouragement to persuade them to visit their GP to be vaccinated. Additionally, family and friends could become involved in attempts to improve vaccine uptake levels by gifting vaccines to their elderly family members [29]. It could be useful if advertising campaigns encouraged the public to offer assistance to transport elderly neighbors and/or relatives to visit their GP to be vaccinated. Lastly, the access issues found in this research appear to indicate that doctors should do their best to offer vaccinations in the course of routine appointments made for other reasons. This could lead to a significant reduction in the number of elderly people with transportation problems having to make an extra journey to the surgery to be vaccinated.

### Limitations and conclusion

This research has the limitation that the study cohort may not be entirely representative of the elderly population as a whole. Recruitment occurred in public places, and so the research cohort may have greater levels of activity in comparison to the wider elderly population. This group (those who were out in public) are worth researching as they have a greater likelihood of being in contact with coronavirus due to their increased outdoors activity, but as participants were most likely to be active elderly persons the research findings are not necessarily applicable to the elderly population as a whole. This undermines our ability to make generalisations from our sample to the population we were studying. Moreover, participants from our study might stay home most of time during the pandemic with limited social interactions. This means that these results cannot necessarily be extrapolated to the general elderly population. Third, the cross-sectional nature of this investigation precludes us from drawing causal inferences. Furthermore, the cross-sectional survey design necessarily represents a snapshot in time, rather than the evolving landscape of the public’s attitudes about COVID-19 vaccination. All the information obtained was self-reported and reporting bias always exists. Although the data was collected from the heterogenous group, we targeted individuals who are willing to participate and give their answers. The individual’s opinion also can be unstable. Any unexpected event could lead to drastic change in their opinion about the vaccination. The final limitation concerns the timing of the survey that might have led to both an overestimate of willingness to receive the vaccination and an underestimate of the vaccine coverage rate among Polish elderly population since the controversy about the efficacy, safety, and necessity of the vaccine against COVID-19. Third wave of COVID-19 pandemic in Poland was growing over the study period [2]. Future longitudinal research is needed to determine the direction of causality for these associations. It would also be desirable to compare the public’s responses in other countries that were similarly affected.

To conclude, this research has demonstrated that current national campaigns and initiatives by healthcare professionals have experienced a high degree of success. Disseminating further information about how effective and safe the vaccine is could lead to even further success. It could also be useful to run advertising campaigns persuading friends and family to actively encourage and assist elderly friends and relatives to accept vaccination.

## Data Availability

Data are available upon request to the corresponding author

## Author Contributions

Conceptualization: MM; methodology: MM and MB; statistical analysis: MM; investigation: MM and MB; writing—original draft preparation: MM; writing—review and editing: MM and MB; funding acquisition: MM. All authors have read and agreed to the published version of the manuscript.

## Institutional Review Board Statement

The project was approved by the local ethics committee of the University of Economics and Social Sciences in Warsaw, Poland.

## Data Availability Statement

Data are available upon request to the corresponding author.

## Footnotes

1 There is a significant gap between intention and actual behavior to get vaccinated. A recent investigation showed that the willingness for influenza vaccination was 45% in general population [20], while the actual vaccination coverage was 9.4%, which was reported by a meta-analysis [21]. Previous studies have examined factors associated with intention of COVID-19 vaccination [7, 9, 10]. However, less is known about COVID-19 vaccination actual uptake and the related factors among older adults. Consequently, for further analysis we included only people who have been already vaccinated and who denied to be vaccinated in the future.

